# Clade-specific multiplex digital PCR assay for monkeypox virus detection in wastewater

**DOI:** 10.1101/2025.02.17.25321728

**Authors:** Melissa Pitton, Charles Gan, Patrick Schmidhalter, Christoph Ort, Timothy R. Julian

## Abstract

Monkeypox virus (MPXV) is a zoonotic pathogen that has recently caused outbreaks in non-endemic areas. Wastewater-based surveillance (WBS) offers substantial potential for monitoring MPVX clades, and it can inform population-level disease dynamics. Here, we report a four-plex digital PCR assay to detect and quantify different clades and subclades of MPXV. The assay demonstrated specificity in distinguishing and quantifying MPXV by clade, which is advantageous for application in both clinical and wastewater settings.

## Background

Mpox is a zoonotic disease caused by the monkeypox virus (MPXV), which was first identified in humans in 1970. Since then, mpox has been endemic in Central and West Africa. Two different clades of MPXV have been described until now, 1 and 2, which are both further classified into subclades (i.e., 1a, 1b, 2a, and 2b). Recent outbreaks in non-endemic regions have raised multiple concerns. Indeed, the World Health Organization (WHO) declared mpox a Public Health Emergency of International Concern twice within a two-year period (2022-2024), which further highlights the importance of monitoring this pathogen.

Diagnostics of MPXV surveillance relies on PCR-based assays for detection and differentiation of MPXV clades. Use of multiplexed qPCR-based assays provides quantitative data on multiple targets when analyzed alongside paired standard curves representing known concentrations, and semi-quantitative data when reporting cycle threshold values alone [1]. Digital PCR (dPCR) is an alternative to qPCR that provides quantification without the need for standard curves. Quantification of MPXV is relevant for assessing viral loads in clinical samples, which can be used to inform disease severity or transmission potential [2, 3]. Quantification is also important for MPXV surveillance within wastewater, which can lead to pathogen detection and improve knowledge about disease dynamics at the population-level over time [4-10]. Importantly, the interim guidance document published by the WHO highlights the potential for wastewater-based surveillance (WBS) to provide actionable information on early detection, differentiation of circulating clades, and quantification of viral loads in sewers which can be used as an indicator for population-level disease dynamics [11].

A dPCR assay that simultaneously allows for clade differentiation is specifically important for WBS, as it can track trends and further highlight the relative proportions of multiple circulating clades within a community. In this study, we adapted four previously published assays to our dPCR platform and then merged the individual assays to develop a four-plex assay for detecting and simultaneously differentiating MPXV clades. The developed assay is intended for applications, which benefit from detection and quantification of individual MPXV clades in clinical or wastewater samples.

### Adapting MPXV assays to our dPCR platform

Four distinct independent assays were first adapted to the Naica dPCR platform from Stilla Technologies (France). These single-plex assays were previously described and validated: G2R_G assay, which targets all clades of MPXV (pan-MPXV) [12]; G2R_WA assay, which is specific to all clade 2 strains (MPXV-Clade2) [12]; C3L assay that specifically targets clade 1a (MPXV-Clade1a) [12]; and dD14-16 assay, which targets the recently emerged clade 1b (MPXV-Clade1b) [13].

Primers and probes are listed and described in Supplementary Table S1 and were manufactured by Microsynth AG (Switzerland). Assays were validated using positive controls consisting of synthetic DNA constructs for each target (i.e., gBlocks®), which were purchased from Integrated DNA Technologies (IDT, USA) (Supplementary Table S2). The assays were developed on the Naica system for Crystal Digital PCR Prism3 or Prism6 (Stilla Technologies), based on channel requirements. All assays were prepared using a total 27 µl pre-reaction volume, which consisted of 5.4 µl of template and 21.6 µl of mastermix. The mastermix was prepared according to the following procedure: PerfeCTa^®^ MultiPlex qPCR ToughMix^®^ (5x) (Cat. No. 733-2324, Quantabio, USA), 0.5 µM of each forward and reverse primer, 0.2 µM of each probe, 0.05 µM of fluorescein, and RNase-free water. The reaction volume (25 μl) was loaded into Sapphire chips. Chips were loaded into a Geode, which partitions the rection mixture into droplets (12 min at 40°C) before thermocycling using the following conditions: enzyme activation (95°C for 10□min), and 45 cycles of denaturation (95°C for 15□s) and annealing/extension (60°C for 1 min).

Each individual target had a specific fluorophore: FAM for pan-MPXV, HEX for MPXV-Clade2, ROX for MPXV-Clade1a, and Cy5 for MPXV-Clade1b. We assessed linearity for each assay by measuring dilution series of positive material in technical replicates. Our data showed strong linearity for each individual assay, with slopes of linear regression ranging from 0.99 to 1.04, and R^2^ values greater than 0.99 (Supplementary Figure S1).

### Development and validation of the novel MPXV four-plex dPCR assay

We developed the four-plex assay by merging the individual assays. The performance of the novel four-plex assay was compared – in parallel – to the one of the single-plex assays. Our findings showed that no difference was observed with respect to quantification, as determined by two-way ANOVA (*p* = 0.74 for pan-MPXV, *p* = 0.05 for MPXV-Clade2, *p* = 0.19 for MPXV-Clade1a, and *p* = 0.59 for MPXV-Clade1b). Since some values were zero, a log transformation was applied using log(concentration + 1). Importantly, all targets originated clusters which were clearly distinguishable, allowing for differentiation between positive and negative partitions, as well as between distinct target genes (Figure 1). Specifically, each target displayed modest to high mean separation scores: 3.9 for pan-MPXV, 2.1 for MPXV-Clade2, 9.8 for MPXV-Clade1a, and 5.7 for MPXV-Clade1b.

**Figure 1.**
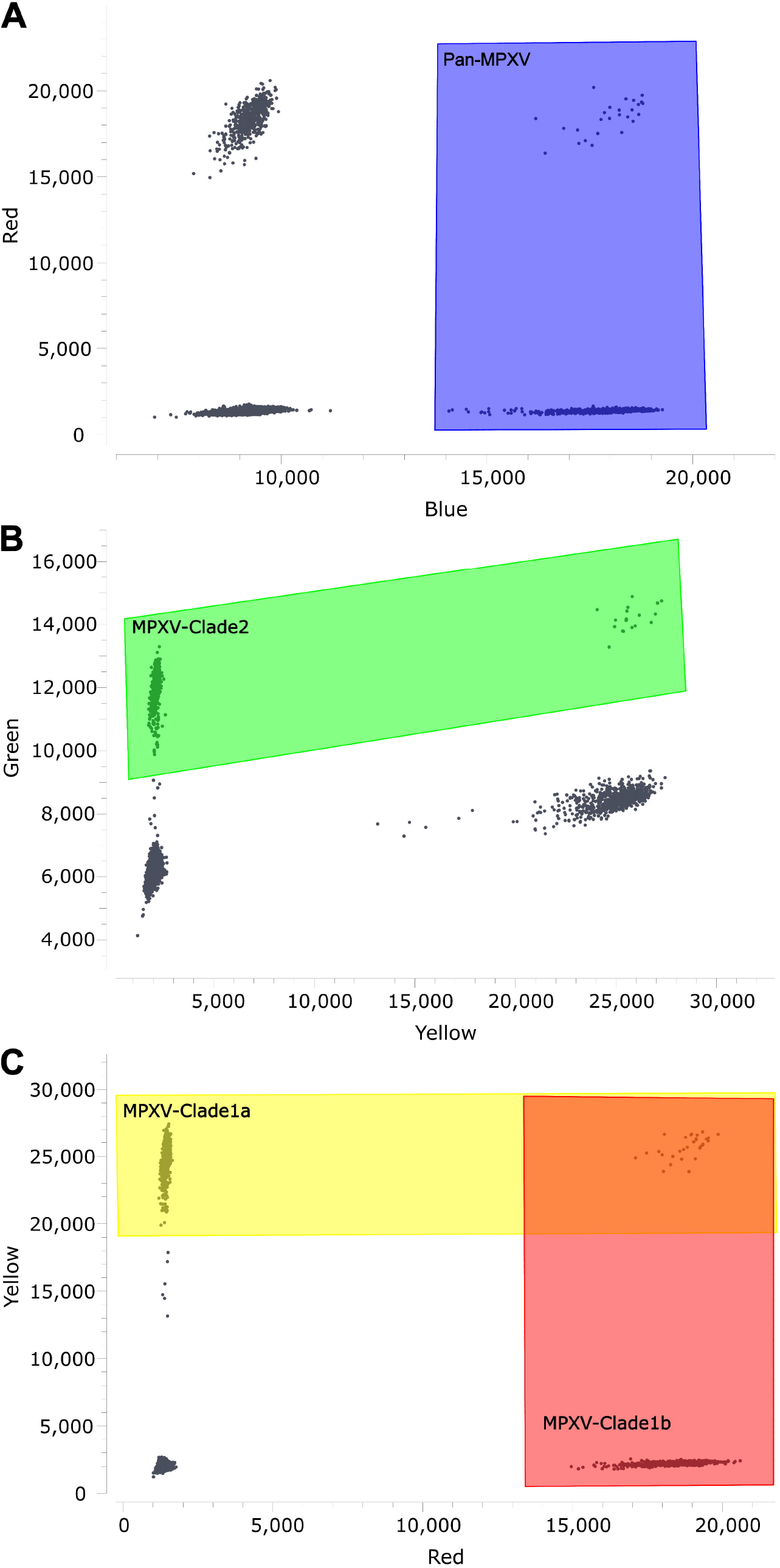
Two-dimensional visualization of the partition classification of a clade-specific dPCR assay targeting MPXV. Both horizontal and vertical axes display fluorescence intensities. Titles on axes indicate the fluorescent channel. Each dPCR partition is represented by a black dot. Positive partitions are separated by negative partitions by manually drawing polygons. Partitions enclosed in the area where polygons overlap are positive for multiple targets (i.e., one partition contains two or more targets). (A) Pan-MPXV positive partitions are separable in the blue channel. (B) MPXV-Clade2 positive cluster can be gated in the green channel. (C) MPXV-Clade1a and MPXV-Clade1b can be gated in yellow and red channels, respectively. This figure, including droplets and colored zones for gating, was generated as a screen shot of the Crystal Miner software (Stilla Technologies), and formatted using Inkscape.

### Sensitivity and specificity of the novel MPXV four-plex dPCR assay

The analytical sensitivity of the assay was assessed by testing three concentrations – 80, 40, and 20 gc/reaction – each measured across a minimum of 15 replicates. The Limit of Detection (LoD) was subsequently determined by probit regression. The LoD at 95% confidence was 97 gc/reaction for MPXV-Clade1a, 93 gc/reaction for MPXV-Clade1b, 85 gc/reaction for MPXV-Clade2, and 51 gc/reaction for pan-MPXV (Supplementary Figure S2). To determine the assay’s specificity, five MPXV clade 2b positive reference samples were tested. All these samples were positive for both pan-MPXV and MPXV-Clade2 targets, and negative for the other two targets, highlighting that the assay could reliably differentiate clade 2 from clades 1a and 1b (Supplementary Figure S3). Importantly, pan-MPXV and MPXV-Clade2 concentrations showed a strong linear relationship (R^2^ > 0.99).

### Proof of concept for wastewater surveillance

Mpox cases have been reported only sporadically in Switzerland since autumn 2022 [14]. Therefore, the assay could not be tested directly on wastewater samples. However, as a proof of concept, synthetic DNA material was spiked into wastewater extracts to determine whether inhibition on PCR amplification is present, and cluster separation is impacted by the wastewater matrix. PCR inhibition was measured as previously described [15], and our analysis showed that mean inhibition was 7.1% for MPXV-Clade1a, 3.0% for MPXV-Clade1b, 8.5% for pan-MPXV, and 3.0% for MPXV-Clade2; which is below our threshold for considering a sample as inhibited (i.e., 40%). Moreover, we observed that the wastewater matrix does not impair the separation score, which describes the discrimination between positive and negative partitions.

## Discussion and conclusion

The application of PCR-based technologies has been proven useful for monitoring MPXV outbreaks. However – in the context of WBS – where different subclades can co-circulate within a community [16], various individual PCR assays need to be implemented for specifically differentiating clades of MPXV. Here, we report the development and validation of a novel four-plex dPCR assay, which can detect MPXV, and differentiate clade 1a and 1b, and clade 2. We validated the assay using synthetic DNA fragments and then addressed the assay’s specificity by applying it on five MPXV clade 2b positive reference samples.

DPCR technology offers absolute quantification of targets without the need of standard curves, which makes it appealing for both wastewater and clinical settings [17], and the multiplexing ability of this assay allows for the simultaneous detection and quantification of multiple circulating clades.

Our study was limited by the absence of MPXV circulation in Swiss wastewater, which prevented the possibility of testing directly the assay on wastewater extracts. However, spiking experiments – performed to mimic wastewater surveillance – showed that the wastewater matrix would not impact the assay’s efficiency, suggesting that the novel multiplex dPCR assay is ready for WBS implementation.

## Data Availability

All data produced in the present study are available upon reasonable request to the authors.

## Acknowledgements

The authors thank the public health laboratory GGD Amsterdam, in particular Dr. Sylvia Bruisten, for kindly providing positive reference samples of monkeypox.

