## Supplemental Material for "Clade-specific multiplex digital PCR assay for monkeypox virus detection in wastewater"

**Supplementary Table S1.** Primers and probes used in the four-plex MPXV assay.

| Assay | Targeted region | Oligonucleotide name | Sequence (5' to 3') | Fluorophore/Quencher | Type | Reference |
|---|---|---|---|---|---|---|
| Pan-MPXV | G2R_G | G2R_G_F | GGAAAATGTAAAGACAACGAATACAG | | Forward primer | Li et al |
| | | G2R_G _R | GCTATCACATAATCTGGAAGCGTA | | Reverse primer | Li et al |
| | | G2R_G _P | AAGCCGTAATCTATGTTGTCTATCGTGTCC | FAM/BHQ-1 | Probe | Li et al |
| MPXV-Clade1a | C3L | C3L_F | TGTCTACCTGGATACAGAAAGCAA | | Forward primer | Li et al |
| | | C3L_R | GGCATCTCCGTTTAATACATTGAT | | Reverse primer | Li et al |
| | | C3L_P | CCCATATATGCTAAATGTACCGGTACCGGA | ROX/BHQ-2 | Probe | Li et al |
| MPXV-Clade1b | dD14-16 | dD14-16_F | AAGACTTCCAAACTTAATCACTCCT | | Forward primer | Schuele et al. |
| | | dD14-16_R | CGTTTGATATAGGATGTGGACATTT | | Reverse primer | Schuele et al. |
| | | dD14-16_P | ATATTCAGGCGCATATCCACCCACGT | Cy5/BHQ-2 | Probe | Schuele et al. |
| MPXV-Clade2 | G2R_WA | G2R_WA_F | CACACCGTCTCTTCCACAGA | | Forward primer | Li et al |
| | | G2R_WA_R | GATACAGGTTAATTTCCACATCG | | Reverse primer | Li et al |
| | | G2R_WA_P | AACCCGTCGTAACCAGCAATACATTT | HEX/BHQ-1 | Probe | Li et al |

**Supplementary Table S2.** Characteristics of synthetic DNA constructs used as positive material in this study.

| Target gene | Type/Description | Sequence (5' to 3') | Source |
|---|---|---|---|
| Pan-MPXV | Synthetic DNA construct (i.e., gBlock®) | ctattattaaatcatgaggtccgtattatactcgtatatattgtttctctcatgtataataataaacggaagagatttagc accacatgcaccatccaatggaaaatgtaaagacaacgaatacagaagccgtaatctatgttgtctatcgtgtcct ccgggaacttacgcttccagattatgtgatagcaagactaatacacaatgtacgccgtgtggttcggatacctttac atctcacaataatcatttacaggcttgtctaagttgtaacggaagatgtgatagtaatcaggtaga | Integrated DNA Technologies (IDT, USA) |
| MPXV-Clade2 | Synthetic DNA construct (i.e., gBlock®) | gaatctgtgaatgctctccaggatattattgtcttctcaaaggagcatcagggtgtagaacatgtatttctaaaacaa agtgtggaataggatacggagtatccggatacacgtctaccggagacgtcatctgttctccgtgtggtcccggaac atattctcacaccgtctcttccacagataaatgcgaacccgtcgtaaccagcaatacatttaactatatcgatgtgg aaattaacctgtatccagtcaacgacacatcgtgtactcggacgaccactaccggtctcagcgaatccatctcaa cgtcggaactaactattaccatgaatcataaagattgtgatcca | Integrated DNA Technologies (IDT, USA) |
| MPXV-Clade1a | Synthetic DNA construct (i.e., gBlock®) | gggaataggatgtgttctatcatactgtactattccgtcacgccccattaatatgaaatttaagaatagtgtagagact gatgctaattacaacataggagacactatagaatatctatgtctacctggatacagaaagcaaaaaatgggacc catatatgctaaatgtaccggtaccggatggacactctttaatcaatgtattaaacggagatgcccatcgcctcgag atatcgataatggccaacttgatattggcggagtagactttggctctagtataacgtactcttgtaatagcggatatca tttgatcggtgaatctaaatcgtattgtgaattaggatcta | Integrated DNA Technologies (IDT, USA) |
| MPXV-Clade1b | Synthetic DNA construct (i.e., gBlock®) | taaaatcagaagttagtacttgtgttaacgatggaaagttatatgtaataggaggattagaattttctatttcaacggg tatagcagaatatttgaaacacggcacttcgaaatggaaaagacttccaaacttaatcactcctagatattcaggc gcatatccacccacgtgtcagattgttaaatgtccacatcctatatcaaacggaaaacttctagcggcttaaaagat catacactcatacaacgacaatgtagactttaagtgcaagtatggatataaactatctggttcctcatcatctacttgc tctccaggaaatacatgacagccggaacttactt | Integrated DNA Technologies (IDT, USA) |

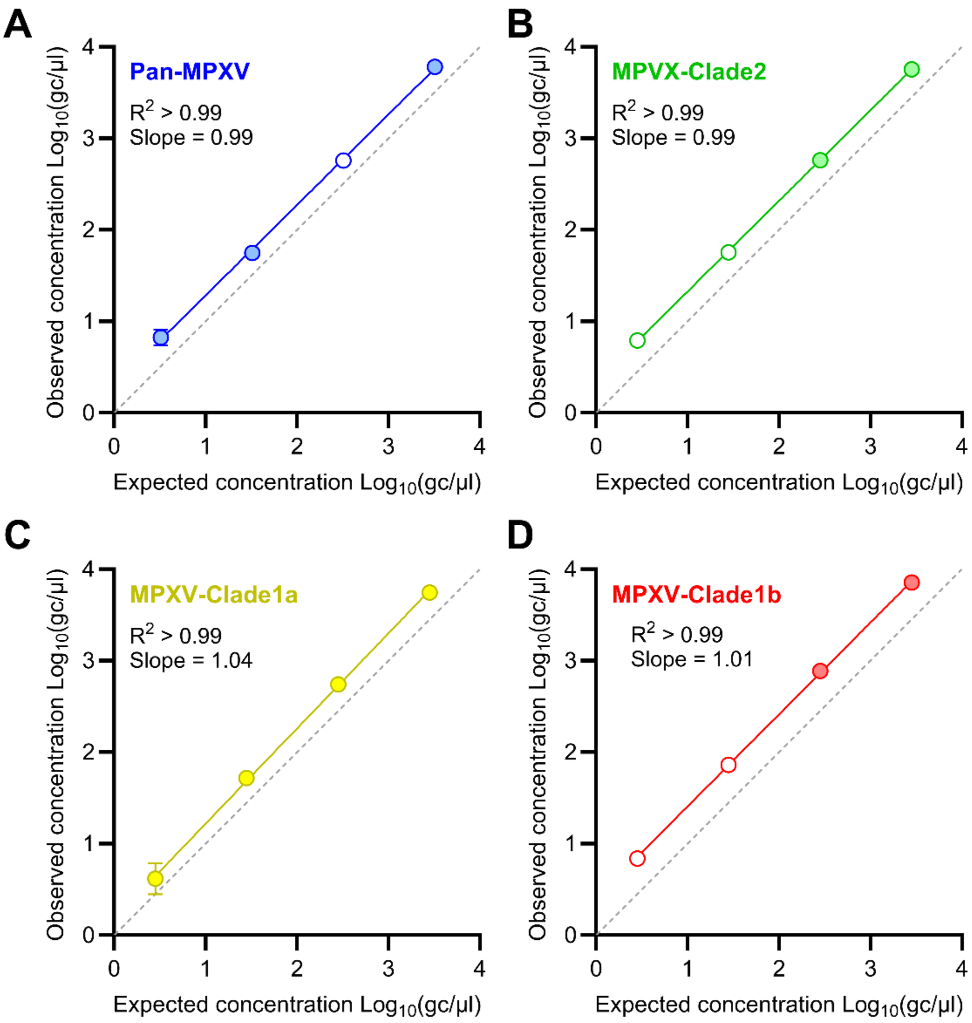


**Supplementary Figure S1. Validation of four individual single-plex assays.** The vertical axes show the observed concentrations of the targets – pan-MPXV in (A), MPXV-Clade2 in (B), MPXV-Clade1a in (C), and MPXV-Clade1b in (D) – measured by dPCR and expressed in log10 genome copies (gc) per microliter (µl) of template. Horizontal axes indicate the expected concentrations provided by the manufacturer. Linear regression lines were fitted to concentrations and $R^2$ values and slopes are displayed on each panel. Error bars indicate the standard deviation around the mean of replicated measurements (n = 2). Empty circles indicate the value of a singlet.

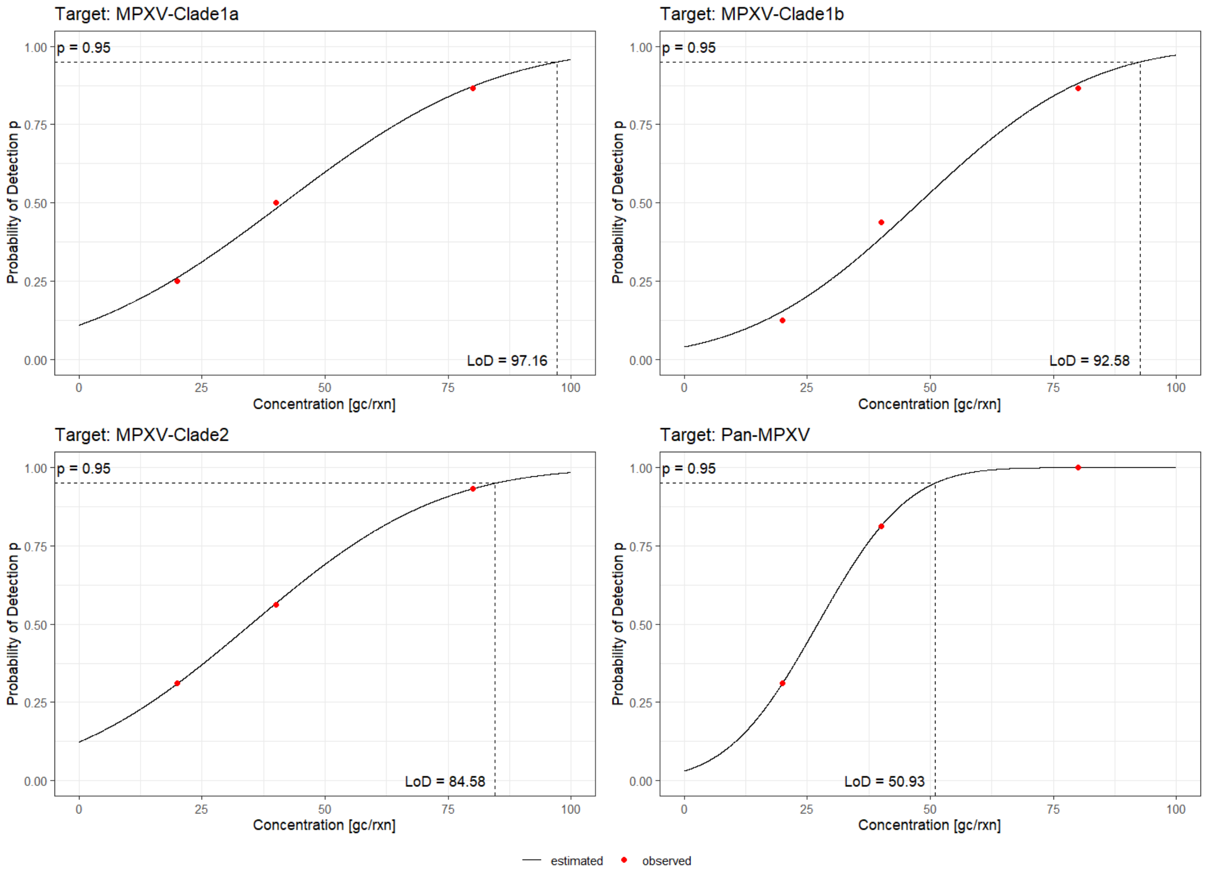


**Supplementary Figure S2. Analytical sensitivity of the novel dPCR assay: limit of detection.** The black curve was modelled using probit regression to fit the probabilities of detection (p) of three concentrations expressed in gene copies per reaction [gc/rxn] (red dots). The horizontal dashed line is set at p = 0.95. The limit of detection (LoD) was determined as the point of intersection of the vertical dashed line with the concentration axis.

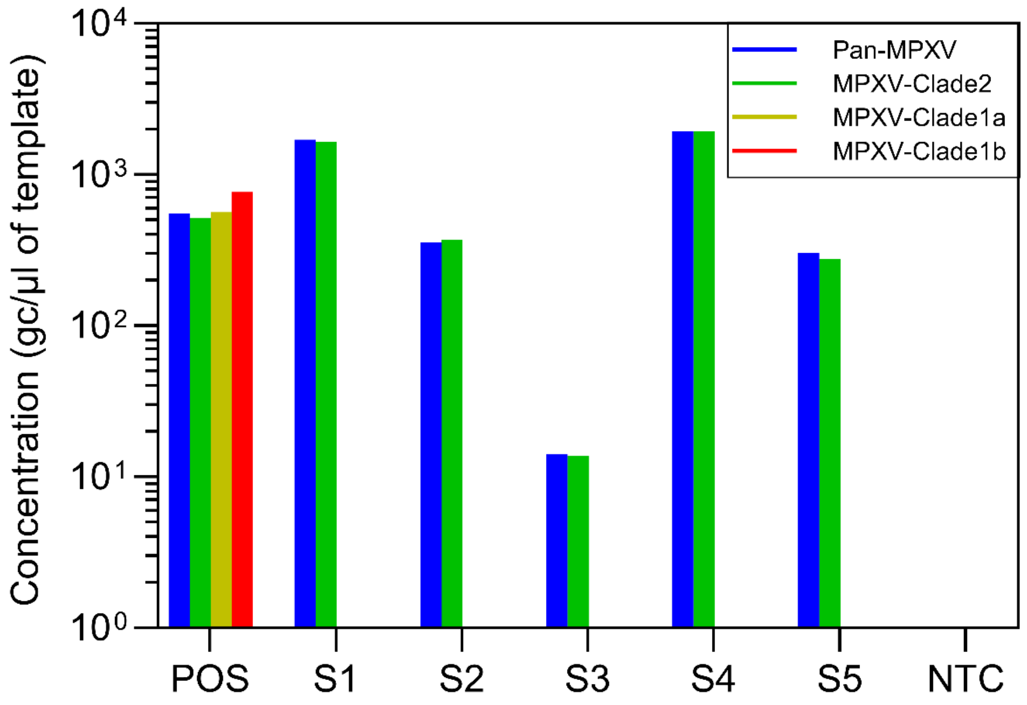


**Supplementary Figure S3. Specificity of the novel four-plex dPCR assay: testing on MPXV positive reference samples belonging to clade 2b.** The horizontal axis indicates the tested samples (S1-S5), as well as the positive control (POS) and the no-template control (NTC). The vertical axis shows the concentration of the four different targets expressed in genome copies (gc) per microliter (µl) of template. Each color represents a target as detailed in the legend. Absence of bars indicates non-detection.
